# Antibiotic Use Among Children Under Two Years With Respiratory Syncytial Virus Infection at Korle Bu Teaching Hospital, Ghana

**DOI:** 10.64898/2026.03.04.26347638

**Authors:** Joycelyn A. Dame, Kwabena A. Osman, An Nguyen, Farina Shaaban, Evangeline Obodai, Clint Pecenka, Louis Bont, Bamenla Goka

## Abstract

**Background:** Respiratory syncytial virus (RSV) is a major cause of lower respiratory tract infections in children, often leading to hospitalisation in infants. In low-resource settings where routine RSV diagnostics are unavailable, clinical overlap with bacterial pneumonia frequently results in unnecessary antibiotic use, contributing to antimicrobial resistance.

**Objective:** To evaluate the frequency and clinical determinants of antibiotic use among RSV-positive children under two years at a tertiary hospital in Ghana.

**Methods:** This cross-sectional study was conducted from June to November 2023 at the Department of Child Health, Korle Bu Teaching Hospital. Children with acute respiratory illness were enrolled and tested for RSV using molecular point-of-care and reverse transcriptase-polymerase chain reaction methods. Antibiotic use and clinical characteristics were analysed among RSV-positive cases.

**Results:** Of 128 children enrolled, 72 (56.2%) tested positive for RSV. Among these, 48 (66.7%) received antibiotics. Antibiotic use was significantly associated with markers of disease severity, including hypoxia (p = 0.009), tachypnea (p = 0.015), dyspnea (p < 0.001), and hospital admission (p < 0.001). Only 11 (23%) had suspected or confirmed bacterial co-infections.

**Conclusion:** A substantial proportion of RSV-positive children received antibiotics. These findings underscore the importance of antimicrobial stewardship programs, rapid diagnostics, and preventive interventions, such as maternal RSV vaccination. Strengthening diagnostic capacity and clinical decision-making in pediatric care is crucial for reducing inappropriate antibiotic use and addressing antimicrobial resistance in low-resource settings.

## Introduction

Respiratory syncytial virus (RSV) is a leading cause of acute respiratory tract infections across all age groups. While it often results in mild symptoms, it can cause severe illness in young children, older adults, and immunocompromised individuals (1). In children, RSV commonly affects the bronchioles, leading to airway obstruction and bronchiolitis (2). The clinical presentation includes fever, cough, tachypnea, increased work of breathing, wheezing, and crackles—symptoms that can overlap with those of other lower respiratory tract infections, such as pneumonia.

The severity of RSV bronchiolitis ranges from mild to severe. Children with mild disease are typically managed at home, while those with moderate to severe symptoms often require hospitalisation for supportive care, including respiratory support(3). In high-resource settings, rapid diagnostic tests are readily available to confirm RSV infection, aiding appropriate clinical decision-making. However, in low-resource settings, such diagnostic tools are often unavailable. As a result, children with viral respiratory infections are frequently treated empirically with antibiotics, especially when hospitalised(4).

While antibiotics may be warranted in critically ill children with suspected bacterial co-infections (5), their routine use in viral infections, such as RSV bronchiolitis, is generally unnecessary and contributes to the growing global threat of antimicrobial resistance (AMR). Inappropriate antibiotic use increases the risk of resistance, which poses significant health and economic burdens. In 2019 alone, bacterial AMR was directly responsible for an estimated 1.27 million deaths and contributed to nearly 4.95 million deaths worldwide(5).

This study aimed to evaluate antibiotic use and its clinical determinants for children presenting with respiratory illness at the Department of Child Health (DCH), Korle Bu Teaching Hospital (KBTH), a tertiary facility in a low-resource setting where RSV testing is not routinely performed.

## Methods

### Study setting

Children included in this cross-sectional analysis were part of the Ghana cohort of the RSV GOLD III Health Economics Study—an international, prospective, multicentre study aimed at quantifying the direct medical, non-medical, and indirect costs associated with RSV infections in both hospitalised and non-hospitalised children under two years of age. The Ghanaian arm of the study was conducted at the DCH in KBTH, Accra, from June to November 2023, a period that typically coincides with the seasonal peak of RSV transmission in the region.

KBTH serves as a major tertiary referral centre for southern Ghana and is the third-largest referral hospital on the African continent, with a capacity of approximately 2,000 beds and 21 clinical and diagnostic departments. DCH provides comprehensive services, including outpatient consultations, emergency care, and inpatient management. Annually, it attends to an estimated 25,000 pediatric patients with a wide range of diverse medical conditions. All pediatric admissions are routed through the department’s emergency unit for triage before onward admission.

Participants for the study were recruited from the outpatient clinic, emergency room, general paediatric wards, and the Paediatric Intensive Care Unit (PICU). Ethical approval was obtained from the KBTH-Institutional Review Board, and written informed consent was obtained from parents or legal guardians prior to enrollment.

### Study population

Children eligible for inclusion in this study were under two years of age and met the World Health Organization (WHO) case definition for (severe) acute respiratory infection ((S)ARI). For outpatient participants, acute respiratory infection (ARI) was defined by the presence of respiratory symptoms—such as cough or shortness of breath—with onset within the preceding 10 days. All those hospitalised were categorised as severe. Infants younger than four days old were excluded, as were children whose clinical presentation was unrelated to respiratory illness or whose respiratory symptoms were attributed to non-infectious causes.

### RSV Diagnosis

Participating children were tested for RSV using a molecular point-of-care (POC) diagnostic device. A nasal swab was taken by a trained research assistant within 72 hours of presenting to the DCH outpatient department, emergency room, PICU or inpatient ward. The sample was tested for RSV using the highly sensitive and specific POC device ID NOW®.

Reverse transcriptase-polymerase chain reaction (RT-PCR) testing of all samples was conducted at the Noguchi Memorial Institute for Medical Research (NMIMR) to confirm the presence of RSV. For this, nasal samples were stored in a transport medium and frozen at −18 °C for later transport on an ice pack to NMIMR. All results were made known to the attending clinicians.

### Data Collection

Eligible participants were enrolled consecutively after their caregivers were informed of the study protocol and provided written informed consent. One eligible participant was not enrolled because the caregiver declined to provide consent. For those who consented, a trained research assistant administered a structured questionnaire to the caregiver, collecting demographic information such as the child’s age, sex, and any known underlying medical conditions.

Clinical information was extracted using a standardised data abstraction form, drawing from the hospital’s electronic medical records. This included details on clinical presentation, antibiotic prescriptions, indications for antibiotic use and number of antibiotics prescribed. Tuberculous drugs, antiretroviral therapy and topical medications were excluded.

Prescribed antibiotics were classified according to the WHO AWaRe categorisation system, which groups antibiotics into Access, Watch, or Reserve categories based on their spectrum, indication, and potential for resistance. All collected data were securely entered into the Castor Electronic Data Capture (EDC) platform.

### Statistical analysis

Data were collected and entered into Excel and Castor files, then merged to Stata dataset for analysis using standard descriptive statistics. Comparisons were made between RSV-positive patients who received antibiotics and those who did not, using the Mann-Whitney test to compare medians and the χ^2^ test to compare categorical variables. A p-value of less than 0.05 was considered statistically significant.

Multivariable logistic regression was performed

## Results

### Clinical Characteristics of Study participants with RSV infection

Between June and November 30, 2023, a total of 129 eligible children were identified, with 128 enrolled after obtaining informed consent. Of these, 72 (56.2%) tested positive for RSV. Fourteen cases initially reported as negative by point-of-care testing were later confirmed positive by RT-PCR at NMIMR and were included as RSV-positive. Two children died, one with RSV and one without. Children with RSV were younger and more likely to present with tachypnea. Among those who tested positive, 48 (66.7%) received antibiotics during the course of their illness.

**Table 1.**
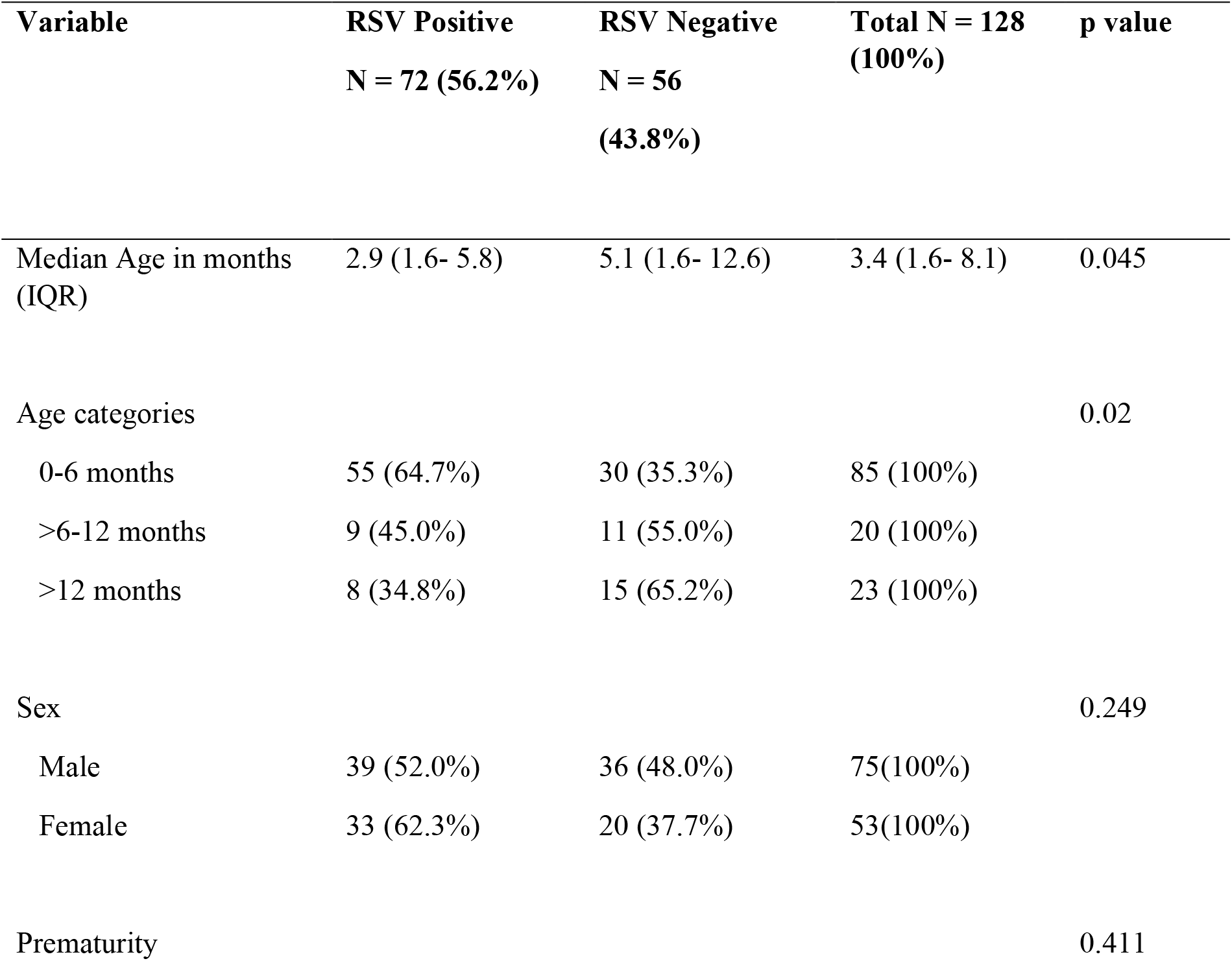
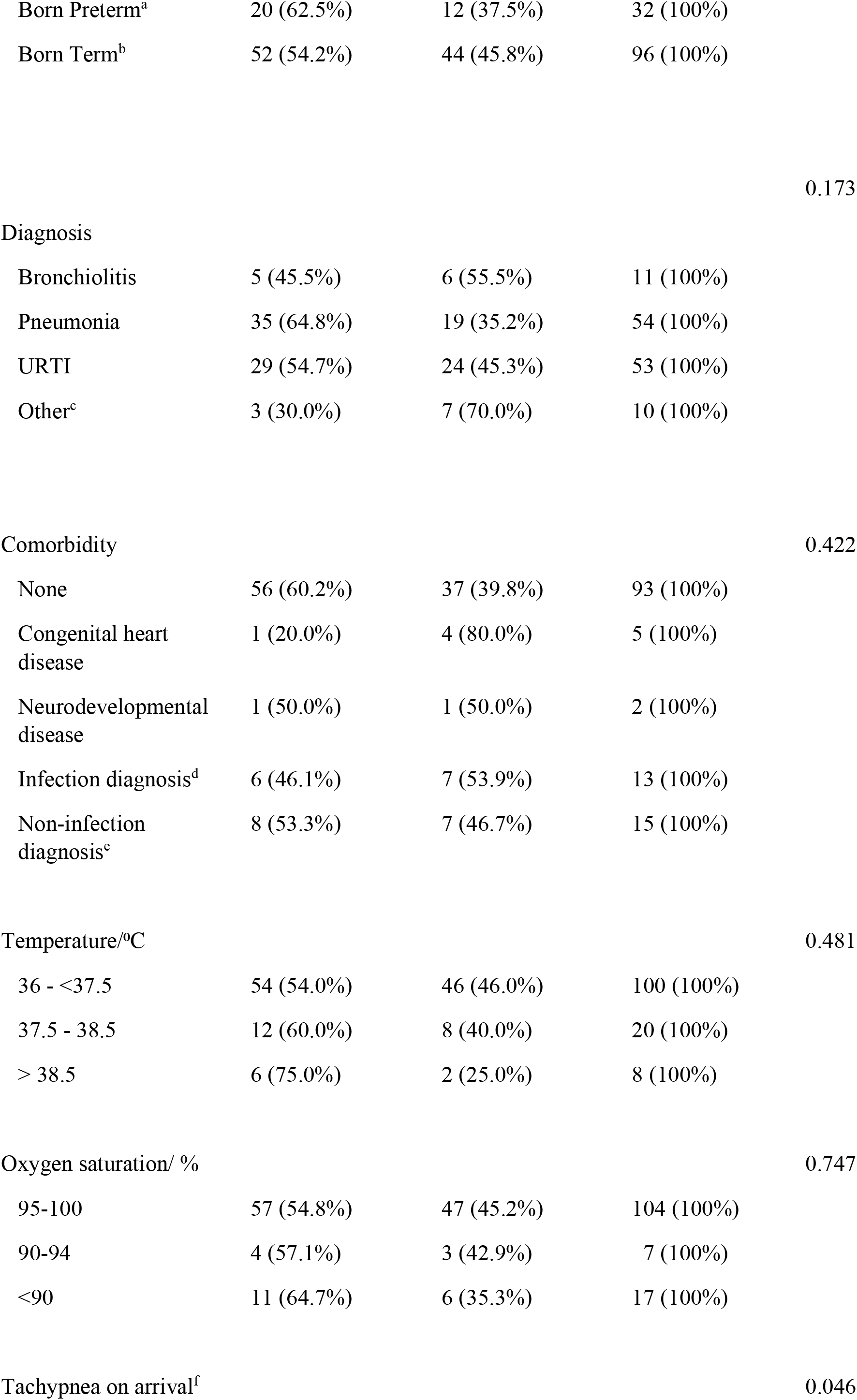
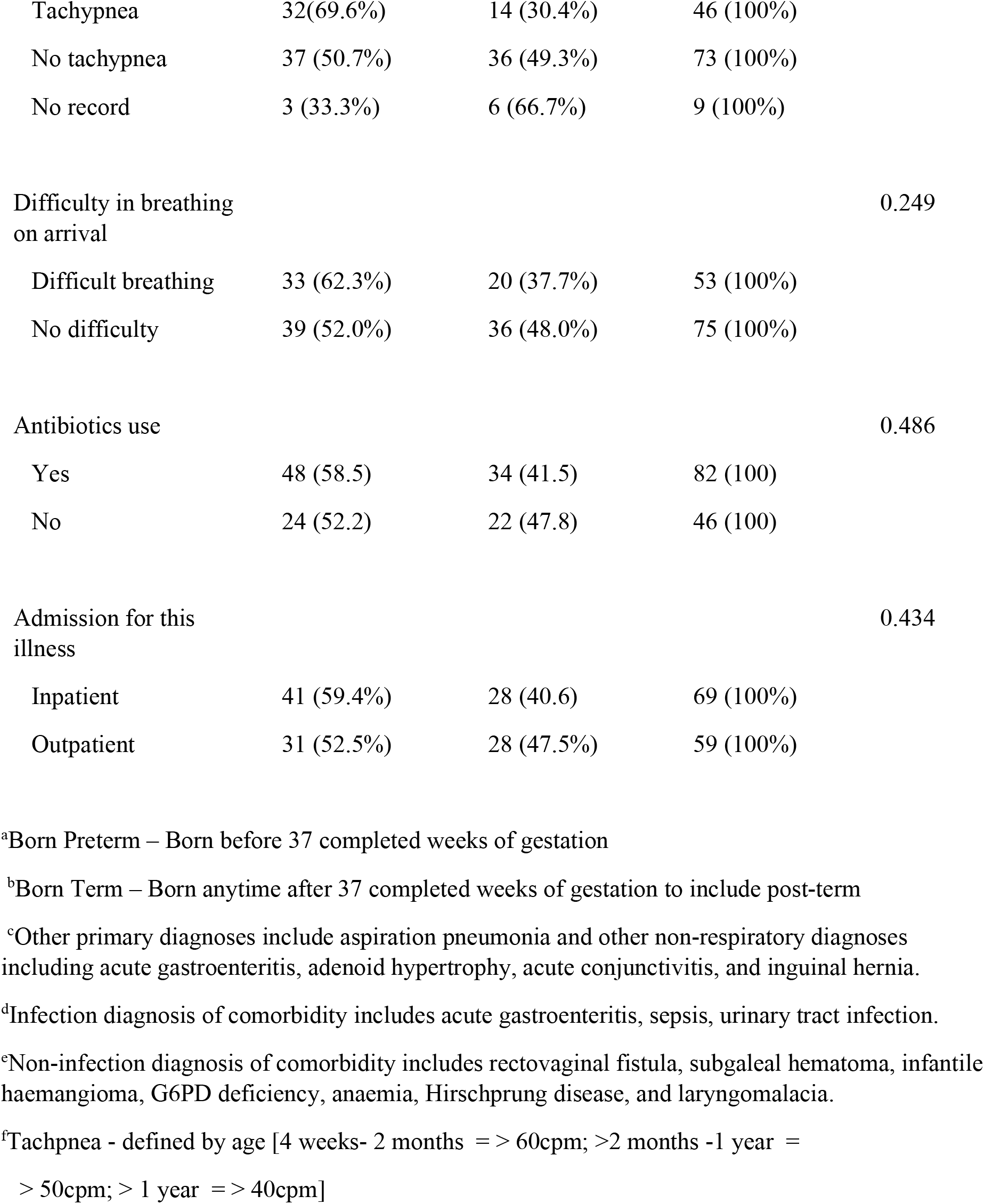
The Clinical Characteristics of Children Categorised by their RSV Diagnosis.

### Clinical Factors Associated with Antibiotic Use Among RSV-Positive Children

Among the 72 children with confirmed RSV infection, 48 (66.7%) received antibiotics during their illness. Children who received antibiotics were more likely to present with lower oxygen saturation (p = 0.009), tachypnea (p = 0.015), difficulty in breathing (p < 0.001), or require hospitalisation (p < 0.001). Antibiotic use was also significantly higher among those who had a suspected or confirmed co-infection (p = 0.129). Those who received antibiotics in this hospitalisation were more likely to be prescribed antibiotics after discharge (p = 0.006). Prior antibiotic use before hospital arrival did not differ significantly between the two groups.

**Table 2.**
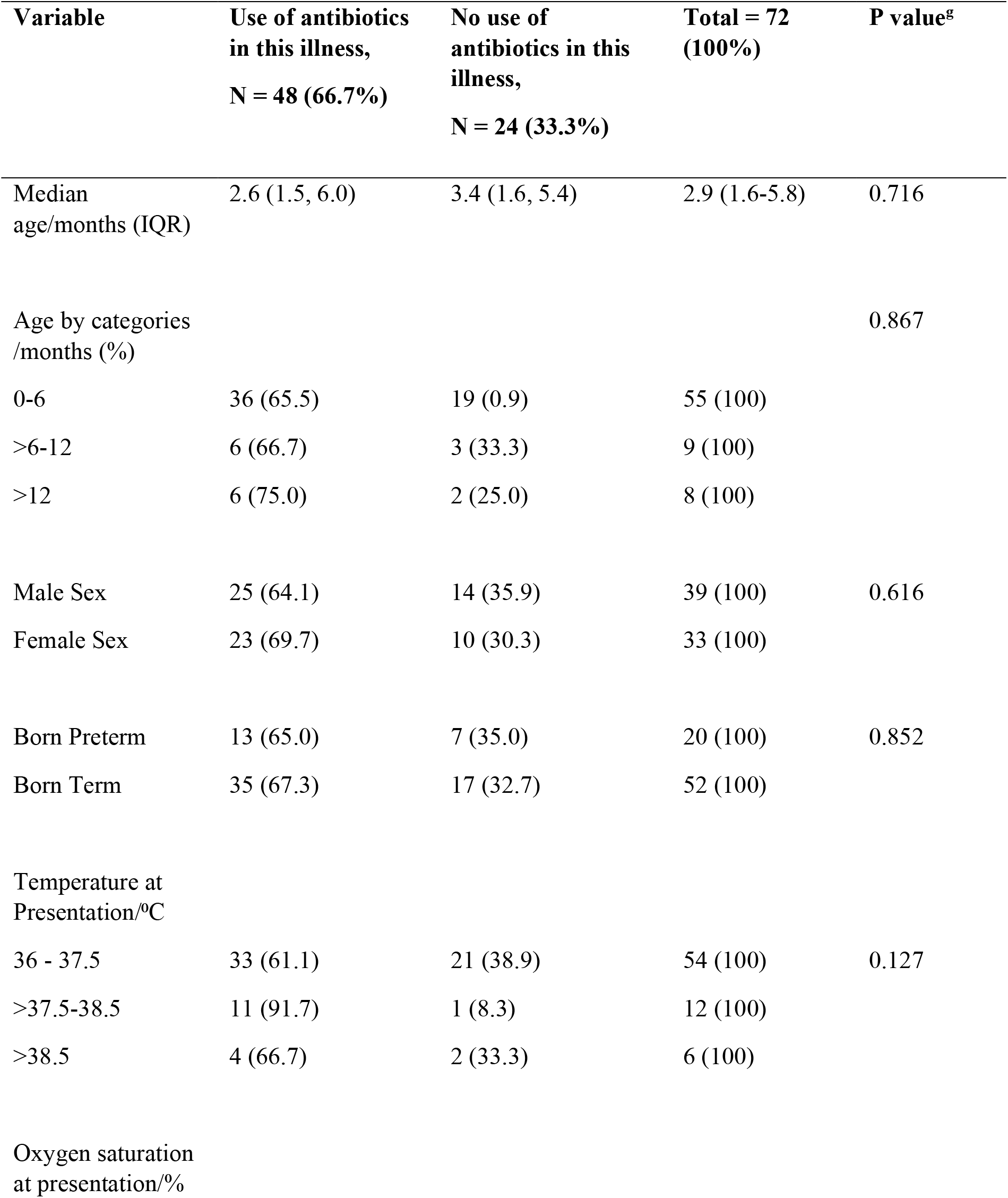
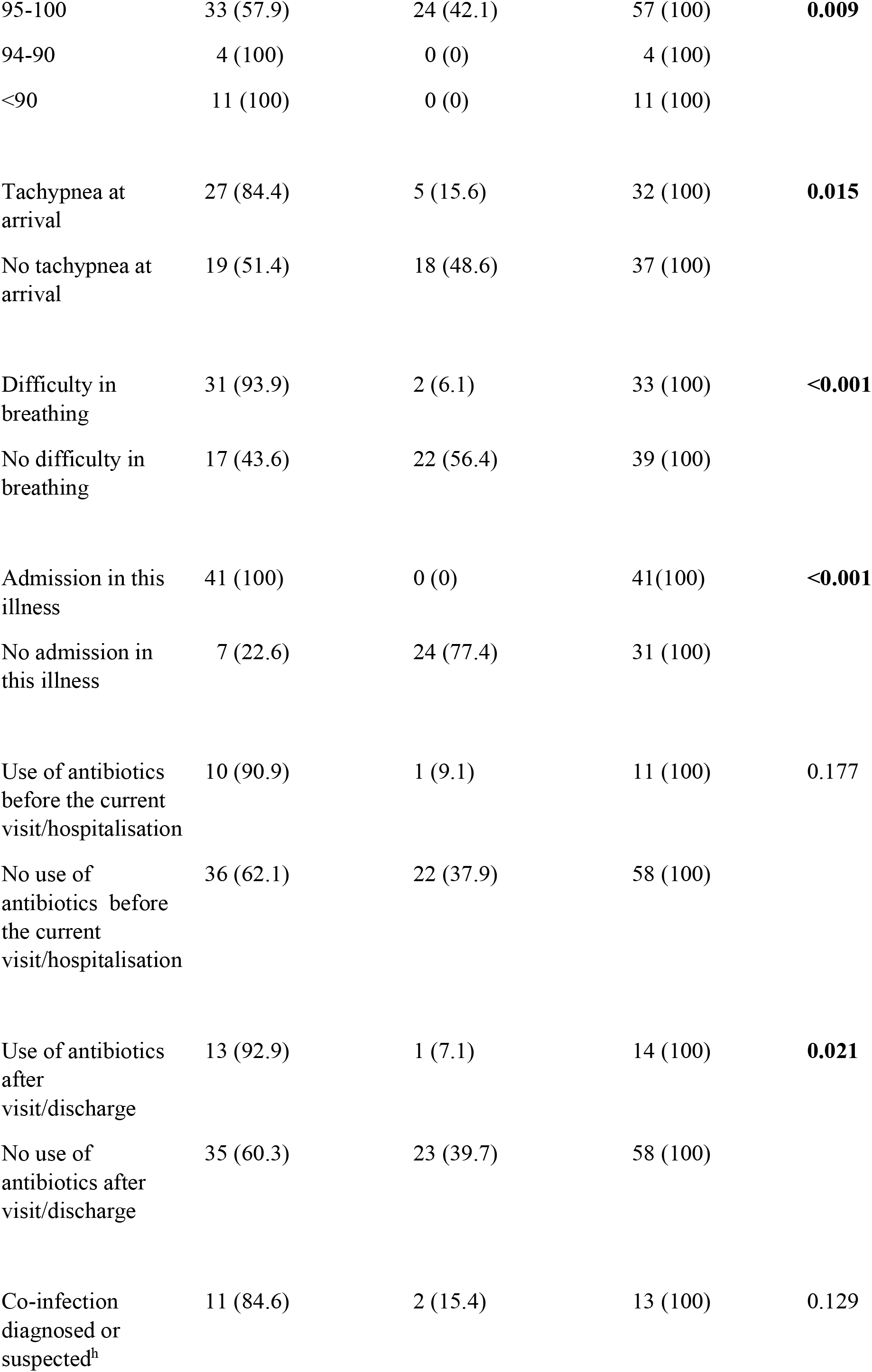
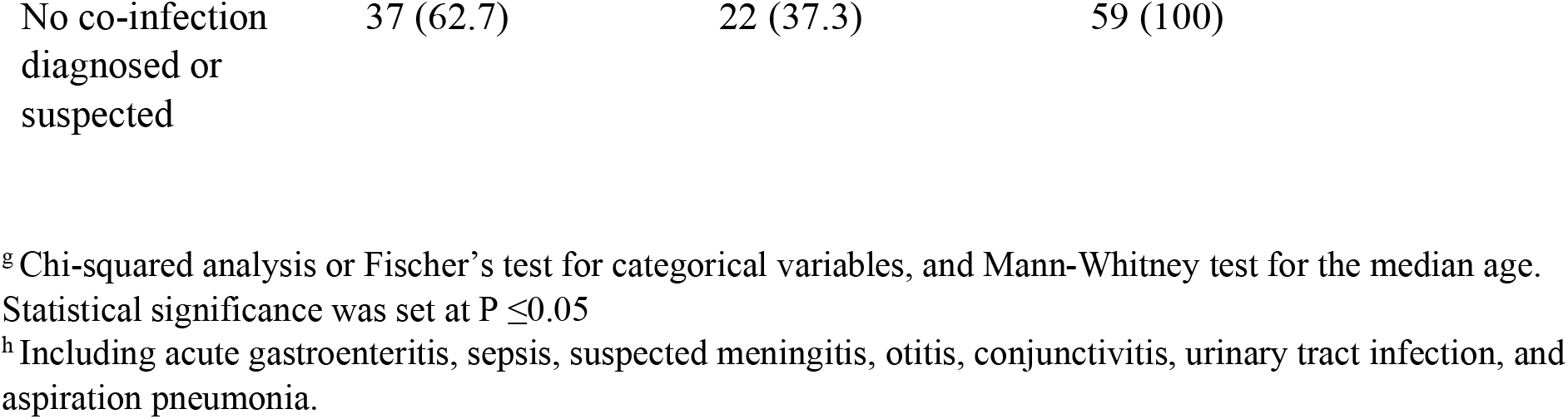
Clinical Factors Associated with Antibiotic Use Among RSV-Positive Children.

| Variable | Use of antibiotics<br>in this illness,<br>N = 48 (66.7%) | No use of<br>antibiotics in this<br>illness,<br>N = 24 (33.3%) | Total = 72<br>(100%) | P value<sup>g</sup> |
| --- | --- | --- | --- | --- |
| Median<br>age/months (IQR) | 2.6 (1.5, 6.0) | 3.4 (1.6, 5.4) | 2.9 (1.6-5.8) | 0.716 |
| Age by categories<br>/months (%) |  |  |  | 0.867 |
| 0-6 | 36 (65.5) | 19 (0.9) | 55 (100) |  |
| >6-12 | 6 (66.7) | 3 (33.3) | 9 (100) |  |
| >12 | 6 (75.0) | 2 (25.0) | 8 (100) |  |
| Male Sex | 25 (64.1) | 14 (35.9) | 39 (100) | 0.616 |
| Female Sex | 23 (69.7) | 10 (30.3) | 33 (100) |  |
| Born Preterm | 13 (65.0) | 7 (35.0) | 20 (100) | 0.852 |
| Born Term | 35 (67.3) | 17 (32.7) | 52 (100) |  |
| Temperature at<br>Presentation/°C |  |  |  |  |
| 36 - 37.5 | 33 (61.1) | 21 (38.9) | 54 (100) | 0.127 |
| >37.5-38.5 | 11 (91.7) | 1 (8.3) | 12 (100) |  |
| >38.5 | 4 (66.7) | 2 (33.3) | 6 (100) |  |
| Oxygen saturation<br>at presentation/% |  |  |  |  |
| 95-100 | 33 (57.9) | 24 (42.1) | 57 (100) | **0.009** |
| 94-90 | 4 (100) | 0 (0) | 4 (100) |  |
| <90 | 11 (100) | 0 (0) | 11 (100) |  |
| Tachypnea at arrival | 27 (84.4) | 5 (15.6) | 32 (100) | **0.015** |
| No tachypnea at arrival | 19 (51.4) | 18 (48.6) | 37 (100) |  |
| Difficulty in breathing | 31 (93.9) | 2 (6.1) | 33 (100) | **<0.001** |
| No difficulty in breathing | 17 (43.6) | 22 (56.4) | 39 (100) |  |
| Admission in this illness | 41 (100) | 0 (0) | 41(100) | **<0.001** |
| No admission in this illness | 7 (22.6) | 24 (77.4) | 31 (100) |  |
| Use of antibiotics before the current visit/hospitalisation | 10 (90.9) | 1 (9.1) | 11 (100) | 0.177 |
| No use of antibiotics before the current visit/hospitalisation | 36 (62.1) | 22 (37.9) | 58 (100) |  |
| Use of antibiotics after visit/discharge | 13 (92.9) | 1 (7.1) | 14 (100) | **0.021** |
| No use of antibiotics after visit/discharge | 35 (60.3) | 23 (39.7) | 58 (100) |  |
| Co-infection diagnosed or suspected<sup>h</sup> | 11 (84.6) | 2 (15.4) | 13 (100) | 0.129 |

|  |  |  |  |
| --- | --- | --- | --- |
| No co-infection<br>diagnosed or<br>suspected | 37 (62.7) | 22 (37.3) | 59 (100) |

<sup>g</sup> Chi-squared analysis or Fischer's test for categorical variables, and Mann-Whitney test for the median age.
Statistical significance was set at $P \leq 0.05$
<sup>h</sup> Including acute gastroenteritis, sepsis, suspected meningitis, otitis, conjunctivitis, urinary tract infection, and aspiration pneumonia.

### Antibiotic Use among 48 Children with RSV infection

Among the 48 RSV-positive children who received antibiotics, a total of 78 antibiotic prescriptions were recorded. The most frequently prescribed class was penicillins, followed by third-generation cephalosporins.

**Figure 1.**
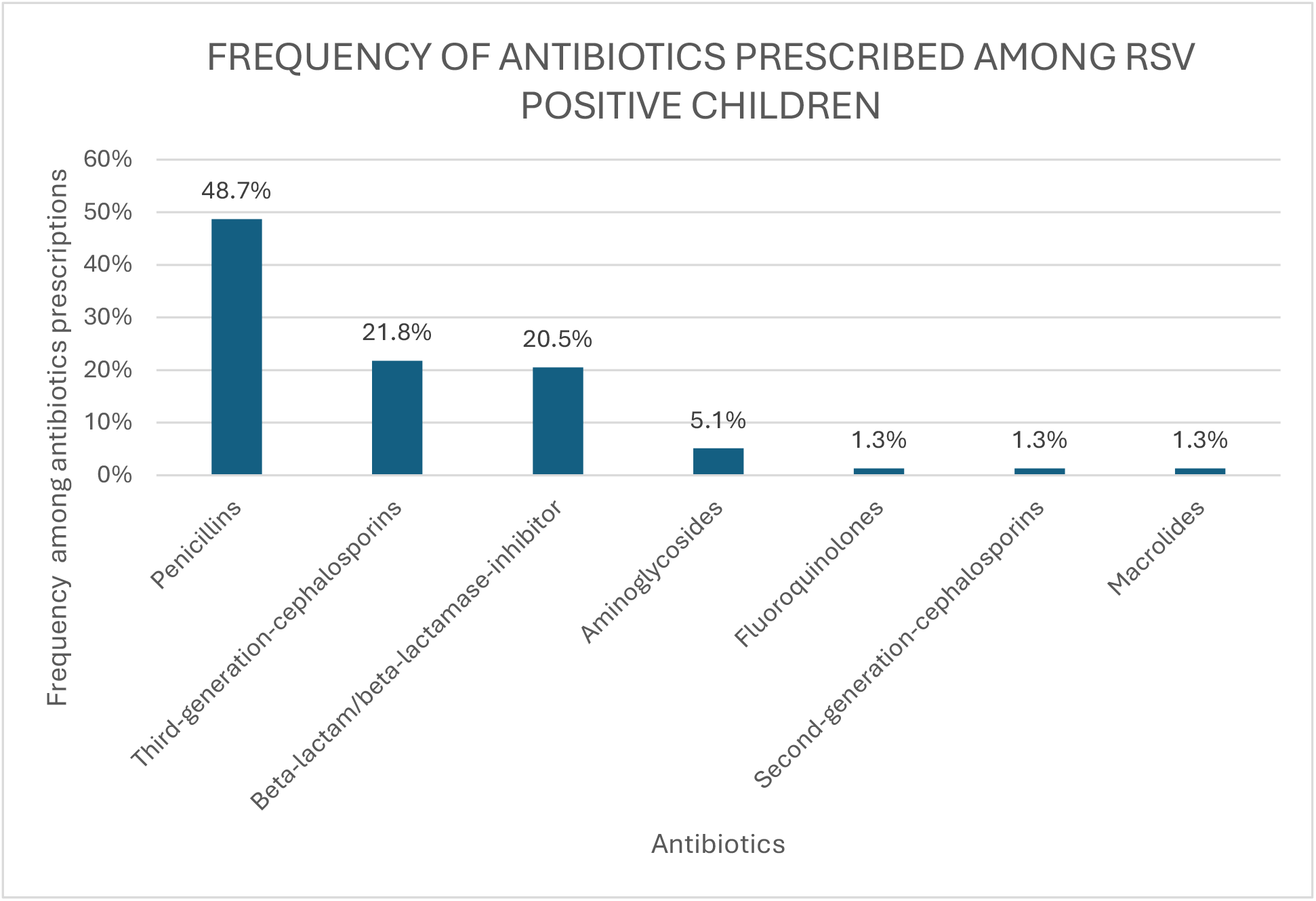
Frequency of antibiotics prescribed among RSV-positive children.

**Figure 2.**
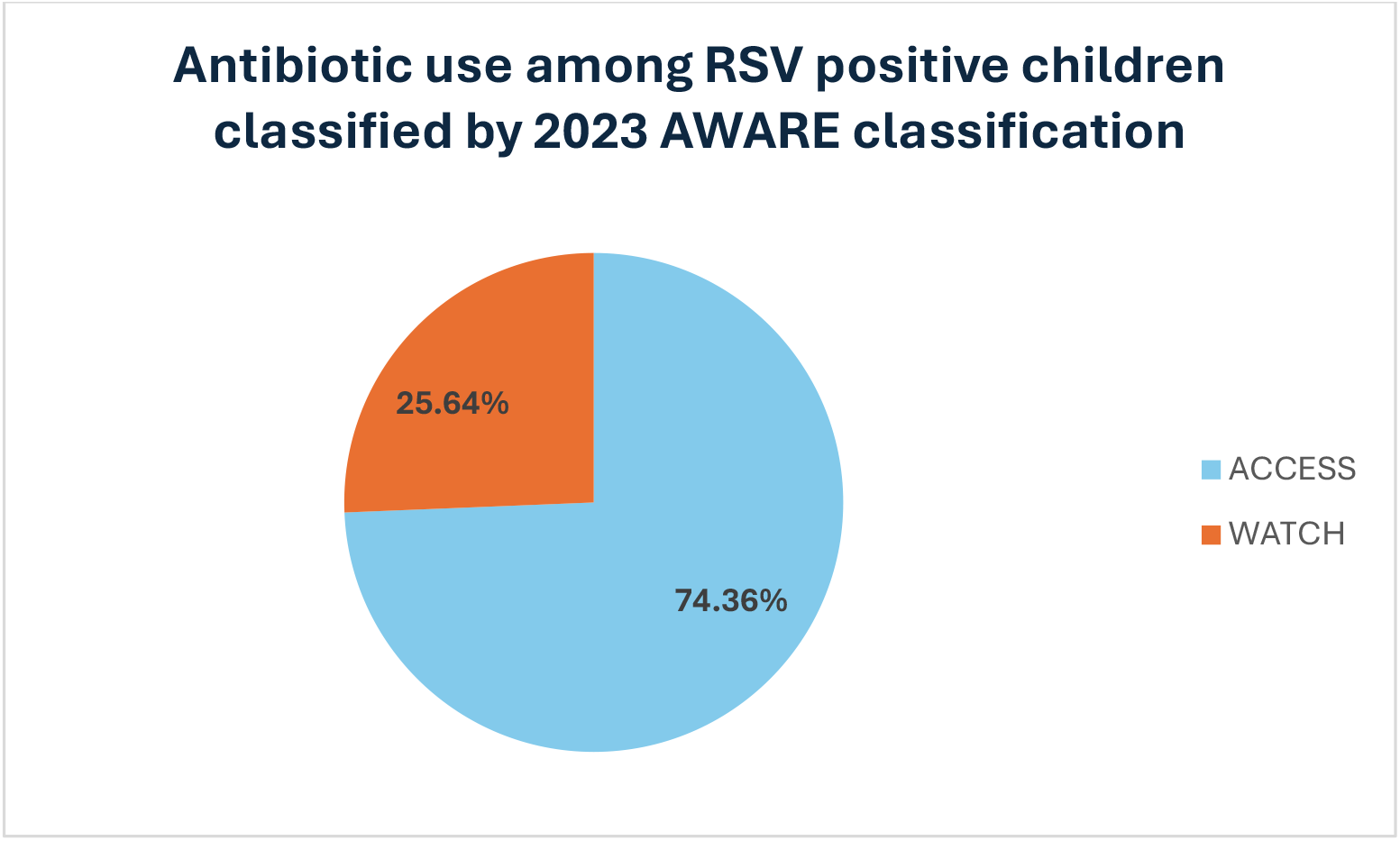
Antibiotics classified by the 2023 AWaRE group of classification

### Diagnoses and co-infection among 48 RSV children who were prescribed antibiotics

Of the children with RSV who received antibiotics, 11 (23%) were diagnosed with either pneumonia or upper respiratory tract infection, together with a co-infection including acute gastroenteritis, urinary tract infection, and sepsis. The remaining 37 children (77%) were diagnosed with pneumonia (n=28), URTI (n=5), and bronchiolitis (n=4) without any co-infection.

**Figure 3.**
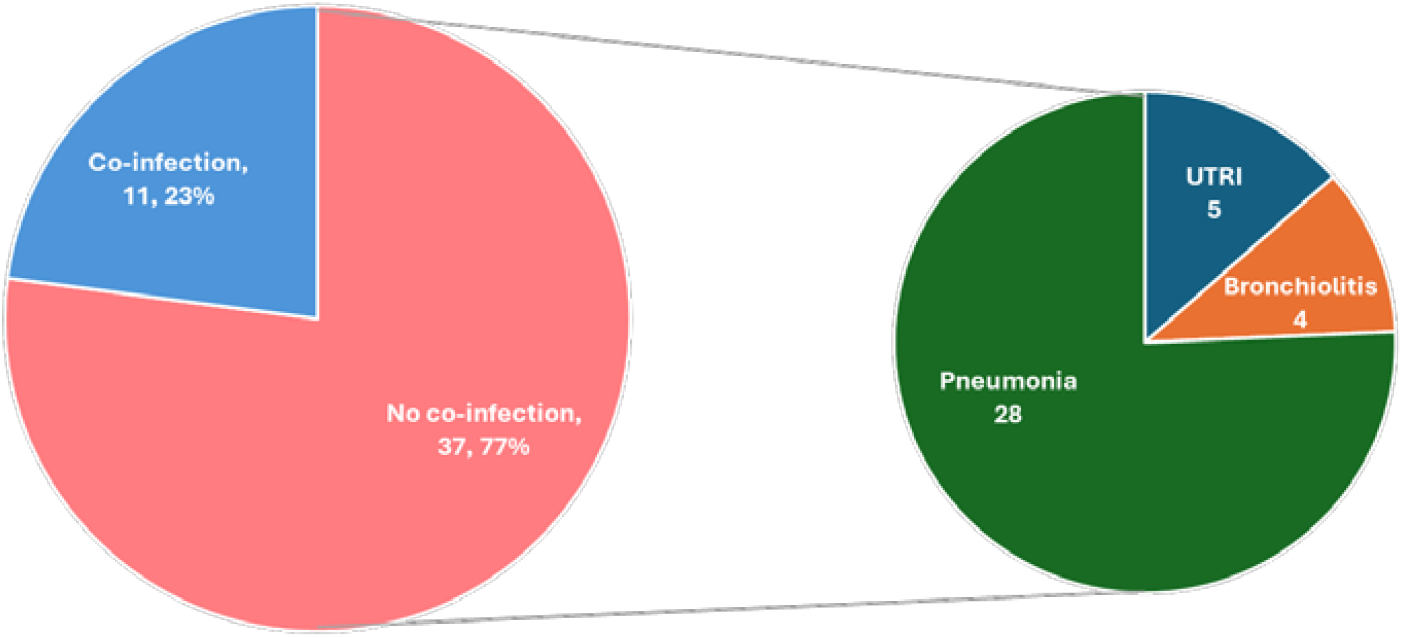
Diagnoses and co-infection among 50 RSV children who were prescribed antibiotics

## Discussion

In this study, nearly 70% of children under two years of age with laboratory-confirmed RSV infection received antibiotics during their illness. This high rate of antibiotic use reflects a common challenge in low-resource settings, where clinical overlap between viral and bacterial respiratory infections, the absence of routine diagnostic tools, and a fear of missing bacterial co-infections often drive empirical antibiotic prescribing. The overuse of antibiotics has important implications for AMR. RSV is a viral illness, and unnecessary antibiotic exposure contributes to resistance by promoting selection pressure on gut and respiratory flora(6). This is particularly concerning in LMICs, where access to second-line antibiotics is limited, and treatment options for resistant infections are few. A systematic review by Luchen et al. of the impact of antibiotics on the gut microbiome of infants in LMICs showed that antibiotics significantly reduce the diversity and alter the composition of the infant gut microbiome in LMICs, while concomitantly selecting for resistance genes whose persistence can last for months following treatment(7).

Our findings underscore the need for the urgent implementation of antimicrobial stewardship programs (ASPs) in pediatric departments, accompanied by clear clinical pathways for managing viral illnesses, such as RSV. These programs should integrate diagnostic testing, reinforce clinical guidelines, train healthcare providers in rational prescribing, and monitor antibiotic use. Investment in such systems is critical to safeguard antibiotic efficacy and improve outcomes in vulnerable pediatric populations.

In this study, antibiotic use was significantly associated with markers of disease severity, including tachypnea, low oxygen saturation, difficulty breathing, and inpatient admission, all of which can mimic bacterial pneumonia. While this may justify the initiation of antibiotics in some cases, our data revealed that 76% of those prescribed antibiotics had no documented bacterial co-infection. Only 24% had a co-diagnosis such as acute gastroenteritis, urinary tract infection, or suspected sepsis. These results suggest a substantial proportion of antibiotics were likely unnecessary, reflecting missed opportunities for targeted prescribing.

Our findings align with previous research. A prospective study conducted in Israel reported that 33.4% of children with confirmed RSV infection but no evidence of bacterial co-infection received unnecessary antibiotic treatment. The likelihood of inappropriate antibiotic use was higher in cases where bacterial cultures were obtained and in children presenting with signs of severe illness, including low oxygen saturation, elevated temperature, rapid breathing, and recent visits to the emergency department (8).

In this study, the use of Watch group antibiotics, particularly third-generation cephalosporins, among RSV-positive children is concerning, given that RSV is a viral illness and does not warrant routine antibiotic therapy, especially not with broad-spectrum agents that carry a higher risk of resistance (6). Thus ASPs that incorporate AWaRe as a core framework for monitoring and optimising pediatric antibiotic use are needed. Implementing hospital-based audit-feedback systems based on AWaRe could reduce the unnecessary use of Watch group antibiotics and support the WHO’s target that at least 80% of national antibiotic consumption should come from the Access group(9).

The availability of a molecular POC test within 24 hours of presentation allowed for the accurate identification of viral aetiology and could have informed antibiotic decision-making. POC testing is increasingly recognised as an essential component of diagnostic stewardship, enabling clinicians to distinguish viral from bacterial infections at the bedside and reduce empirical antibiotic use (10). A randomised controlled trial in Vietnam demonstrated that C-reactive protein-guided antibiotic prescribing, when combined with viral testing, resulted in a reduction of over 40% in antibiotic use among children with respiratory illnesses (11). Implementing similar stewardship strategies, including syndromic POC testing, could improve prescribing practices at KBTH and other tertiary centres in Ghana.

Importantly, these findings also strengthen the case for RSV prevention through maternal immunisation and monoclonal antibodies in infants. The WHO recommends that all countries introduce RSV immunisation products to prevent severe disease in infants, using either maternal vaccination (RSVPreF) or infant monoclonal antibody (nirsevimab), depending on local context. Countries are to consider factors such as cost, supply, health system readiness, and implementation feasibility when choosing between the two products(12). By preventing severe illness, RSV immunisation can reduce unnecessary antibiotic use, supporting its inclusion in national immunisation programs as a strategy to combat AMR. Our findings reinforce this approach, particularly in settings with a high RSV burden and frequent empirical antibiotic prescribing.

### Limitations

While RSV was confirmed using molecular POC and PCR testing, routine bacterial testing was not performed, potentially underestimating appropriate antibiotic use. Additionally, the rationale behind antibiotic prescribing was not explored, limiting insight into decision-making under diagnostic uncertainty. As a single-centre study conducted at a large urban tertiary hospital, generalizability to smaller or rural settings may be limited. Lastly, long-term outcomes such as length of stay or treatment response were not assessed.

### Conclusion

This study demonstrated a high prevalence of antibiotic use in RSV-positive children under two years, primarily driven by clinical severity and diagnostic uncertainty. These findings highlight the critical need to incorporate rapid diagnostics, antimicrobial stewardship programs ASPs, and preventive measures such as maternal RSV vaccination into paediatric care to reduce inappropriate antibiotic use and help combat AMR. Tackling these challenges will require coordinated efforts across clinical practice, diagnostic capacity, and health policy in Ghana and other low-resource settings.

## Data Availability

All data produced in the present study are available upon reasonable request to the authors

## References

1. Munro AP, Martinón-Torres F, Drysdale SB, Faust SN. The disease burden of respiratory syncytial virus in Infants. Current opinion in infectious diseases. 2023;36(5):379–84.

2. Florin TA, Plint AC, Zorc JJ. Viral bronchiolitis. The Lancet. 2017;389(10065):211–24.

3. Ajayi OO, Ajufo A, Ekpa QL, Alabi PO, Babalola F, Omar ZT, et al. Evaluation of bronchiolitis in the pediatric population in the United States of America and Canada: a ten-year review. Cureus. 2023;15(8).

4. Machowska A, Stålsby Lundborg C. Drivers of irrational use of antibiotics in Europe. International journal of environmental research and public health. 2019;16(1):27.

5. Murray CJ, Ikuta KS, Sharara F, Swetschinski L, Aguilar GR, Gray A, et al. Global burden of bacterial antimicrobial resistance in 2019: a systematic analysis. The lancet. 2022;399(10325):629–55.

6. Laxminarayan R, Matsoso P, Pant S, Brower C, Røttingen J-A, Klugman K, et al. Access to effective antimicrobials: a worldwide challenge. The Lancet. 2016;387(10014):168–75.

7. Luchen CC, Chibuye M, Spijker R, Simuyandi M, Chisenga C, Bosomprah S, et al. Impact of antibiotics on gut microbiome composition and resistome in the first years of life in low-to middle-income countries: A systematic review. PLoS Medicine. 2023;20(6):e1004235.

8. Obolski U, Kassem E, Na’amnih W, Tannous S, Kagan V, Muhsen K. Unnecessary antibiotic treatment of children hospitalised with respiratory syncytial virus (RSV) bronchiolitis: risk factors and prescription patterns. Journal of Global Antimicrobial Resistance. 2021;27:303–8.

9. Zanichelli V, Sharland M, Cappello B, Moja L, Getahun H, Pessoa-Silva C, et al. The WHO AWaRe (Access, Watch, Reserve) antibiotic book and prevention of antimicrobial resistance. Bulletin of the World Health Organization. 2023;101(4):290.

10. Moore LS, Villegas MV, Wenzler E, Rawson TM, Oladele RO, Doi Y, et al. Rapid diagnostic test value and implementation in antimicrobial stewardship across low-to-middle and high-income countries: a mixed-methods review. Infectious Diseases and Therapy. 2023;12(6):1445–63.

11. Do NT, Ta NT, Tran NT, Than HM, Vu BT, Hoang LB, et al. Point-of-care C-reactive protein testing to reduce inappropriate use of antibiotics for non-severe acute respiratory infections in Vietnamese primary health care: a randomised controlled trial. The Lancet Global Health. 2016;4(9):e633–e41.

12. World Health Organization. Summary of WHO Position Paper on Respiratory Syncytial Virus (RSV) Immunization 2025.Date of access 06 July 2025, https://cdn.who.int/media/docs/default-source/immunization/position_paper_documents/hpv/summary_respiratory_syncytial.pdf?sfvrsn=e

